# COVID-19 isolation and containment strategies for ships: Lessons from the USS Theodore Roosevelt outbreak

**DOI:** 10.1101/2020.11.05.20226712

**Authors:** Madison Stoddard, Kaitlyn Johnson, Doug White, Ryan Nolan, Natasha Hochberg, Arijit Chakravarty

## Abstract

The control of shipborne disease outbreaks represents a vexing but entirely predictable challenge at the start of any pandemic. Passenger ships, with large numbers of people confined in close quarters, can serve as incubators of disease, seeding the pandemic across the globe as infected passengers return home. Short-term steps taken by local authorities can exacerbate this problem, creating humanitarian crises and worsening the scale of the outbreak. In this work, we have undertaken a model-based examination of the USS Theodore Roosevelt outbreak to understand the dynamics of COVID-19 spread aboard the aircraft carrier. We have used a series of counterfactual “what-if” analyses to better understand the options available to public health authorities in such situations. The models suggest that rapid mass evacuation and widespread surveillance testing can be effective in these settings. Our results lead to a set of generalizable recommendations for disease control that are broadly applicable to the current COVID-19 crisis as well as to future pandemics.

## Introduction

Congregate settings such as nursing homes, schools and passenger ships have posed a particular problem in the COVID-19 pandemic (Moriarty, 2020). The disease burns quickly through these high-contact populations, and such outbreaks have been widely publicized (Farber, 2020; Taylor, 2020; Van Beusekom, 2020) and in some cases deadly (Cain, 2020).

Shipborne outbreaks have proven especially challenging to contain. The availability of disease detection tests may be limited on board, and increasing testing capacity may require additional medical staff and air-drop of testing materials (Yamahata and Shibata, 2020). Making matters worse, test processing facilities may not be available on board, resulting in slow turnaround times for test results (Dyer, 2020). Most passenger ships have very limited capacity for isolating infected passengers (either via airlift or onboard isolation), and in the early days of the COVID-19 pandemic, mass evacuation of vessels bearing infected passengers was often blocked at port by local authorities (Gilday, 2020; Yamahata and Shibata, 2020). This problem is compounded aboard military vessels where the entire crew is placed in very close quarters, and fan-forced ventilation recirculates spent and unfiltered air (LaGrone, Apr 2020; Malone, 2020). The high degree of contact among all passengers aboard a typical ship facilitates spread and renders contact tracing effectively impossible (Crozier, 2020).

The ramifications of at-sea outbreaks have been broad, impacting the cruise industry and the military in high-profile events such as the Diamond Princess cruise ship and USS Theodore Roosevelt aircraft carrier outbreaks in early 2020 (Mallapaty, 2020). A precise quantitative understanding of the dynamics of transmission in such a setting can be invaluable in formulating strategies for the control of future outbreaks, both for COVID-19 and for other pandemics. The dataset for the USS Theodore Roosevelt (CVN-71, referred to in this paper as “the Roosevelt”) provides detailed insight into the kinetics of shipborne outbreaks, thanks to the Navy’s record-keeping and transparency. In this paper, we examined the Roosevelt outbreak closely through a quantitative lens to evaluate the relative effectiveness of different disease control strategies.

The Roosevelt outbreak resulted in the death of one sailor, several hospitalizations, extensive negative press, significant costs for on-shore isolation facilities, and an approximately two-month departure from duty for the aircraft carrier (LaGrone, Jun 2020; Peniston, 2020). These negative outcomes were aggravated by the lack of FDA-approved SARS-CoV-2 molecular tests at the time. Notably, these outcomes occurred in spite of the high degree of compliance and top-down command of the Navy, an open airbridge to shore-based facilities at all times during the outbreak based on the carrier’s own air wing (“USNI News Fleet and Marine Tracker,” 2020), and access to a mobile laboratory capable of testing for SARS-CoV-2 while at sea (Peniston, 2020; “USNI News Fleet and Marine Tracker,” 2020).

The Navy’s approach to containing the outbreak on the Roosevelt involved testing-based isolation and mass evacuation to quarantine facilities on Guam. The first case was detected on March 22, 2020 while the carrier was underway in the Philippine Sea (“USNI News Fleet and Marine Tracker,” 2020), so mass evacuation was delayed until the ship reached Guam (at the edge of the Philippine Sea, and in theory, less than two days’ sail away for a carrier strike group (Brain, 2004)). Sailors testing positive before this time were either airlifted to land or quarantined in the limited isolation facilities aboard (Gilday, 2020). The Roosevelt docked in Guam on March 27^th^, with the crew being restricted to the ship and the pier. Mass evacuation started on March 31st, and crewmembers were evacuated until data collection stopped on May 5, at which point 796 sailors remained on board and 1,156 cases (23.8%) had been documented among the crew (LaGrone, Jun 2020). In a serology study performed by the CDC, a 60% incidence of SARS-CoV-2-reactive antibodies was observed. The study cohort was biased in favor of crewmembers having previously tested positive, with 235 of 382 volunteers (61%) having previously received a positive test result by RT-PCR (Payne, 2020). This result is consistent with a high degree of detection accuracy by the end of the Roosevelt outbreak.

In the present investigation, we leverage publicly available data from the Roosevelt outbreak to fit an epidemiological model and test strategies for outbreak mitigation. We evaluate the effectiveness of the Navy’s disease containment strategy and provide recommendations for high-contact at-sea settings with an emphasis on naval readiness. Our findings also provide insight more generally into strategies for the control of COVID-19 and other infectious diseases in shipborne settings.

## Methods

### Data collection

Data regarding cumulative cases of SARS-CoV-2 infection by date among the Roosevelt crew were compiled from publicly available military and civilian sources (Dyer, 2020; LaGrone, 2020, p. 600, 2020; Peniston, 2020). Cases were detected by molecular testing and do not include serological positives. Case counts include sailors who tested positive for COVID-19 while in isolation on Guam after evacuation from the ship, implying exposure while on board. When available, we also compiled data describing the number of crewmembers remaining aboard over time to reflect the Navy’s evacuation of the ship. This data is summarized in Figure 1A.

**Fig 1.**
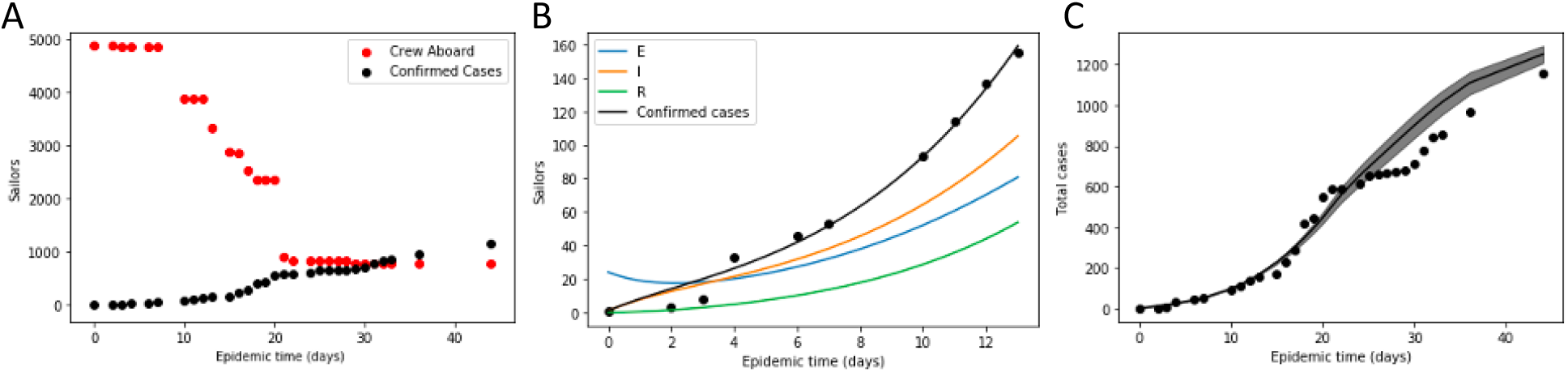
SEIR model describes disease spread in the Roosevelt outbreak. A) Crew aboard and confirmed cases on the Roosevelt throughout the duration of the outbreak. B) SEIR model fit to the Roosevelt case data for the first two weeks of the outbreak (E, I, and R compartments shown). Lines represent model fit, while black dots represent confirmed case data. C) 95% confidence interval for the model predicted total cases assuming random evacuation of sailors (black shaded region) compared to reported case counts (black dots).

## SEIR model fit to early outbreak dynamics

To determine the population-specific epidemiological parameters for SARS-CoV-2 spread in the Roosevelt outbreak, we fit an ordinary differential equation (ODE)-based compartmental model to the cumulative caseload data for the outbreak. The population is partitioned into four compartments representing susceptible (S), exposed (E), infectious (I), and recovered (R) individuals. We leveraged disease-specific model parameters—the latency and recovery period—from studies in the literature leveraging richer datasets describing more complete outbreak time courses, which are better suited to this purpose (Rocklöv et al., 2020). The population-specific parameters—the contact period and the initial exposed population—were fitted to the cumulative detected case data.

Equations 1-5: ODEs for pre-intervention SEIR model

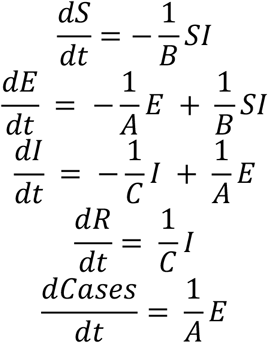

Initial conditions for the SEIR model

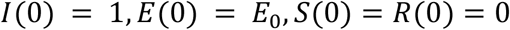

The latency period A in the SEIR model is the duration of pre-infectious exposure. The recovery period C is the duration of infectiousness or the time to recovery. These two parameters are disease-specific and do not vary based on behavior. The contact period B describes the average time to transmission for an infected person and is dependent on the contact behaviors of the population. R_0_, the intrinsic reproductive number in a population, is the ratio of C/B (Ridenhour et al., 2014). The initial exposed population E_0_ is the predicted number of exposed sailors at the time of the first detected case. The model is agnostic to the source of these exposures.

We used a least squares minimization to fit the model-predicted number of infections to the reported cases from the first two weeks of the outbreak. We assumed disease dynamics in this period reflect the underlying rate of disease spread before outbreak response measures such as increased testing-based isolation and mass evacuation began impacting the number of cases. These fit parameters do not represent truly unchecked spread, as basic pandemic preparedness measures such as enhanced cleaning, limited testing, and symptom-based isolation were already in place on the Roosevelt when the outbreak emerged (Gilday, 2020). We assume the contact rate did not change throughout the duration of the outbreak and we assume there are no additional introductions of disease from outside the ship. Likewise, we used this fit contact rate to inform the modeling implemented in the remainder of our analysis.

### Model-based assessment of sailor removal strategies for outbreak containment

The Navy employed two strategies to control the Roosevelt outbreak: removal of sailors at random by mass evacuation and targeted removal of sailors who tested positive by isolation (either onboard or onshore) (LaGrone, Jun 2020). To evaluate and optimize these strategies in the context of shipborne outbreaks, we varied the key implementation parameters for each strategy and the combination of both strategies. The testing-based isolation strategy is defined by the percent of the initial crew tested each day, which is assumed to be constant throughout the outbreak. For the evacuation-based strategy, we explore the impact of evacuation rate, defined as the percent of the initial crew removed each day, the final crew size at which evacuation is stopped, and the timing of the start of evacuation. We assume that the underlying epidemiological parameters – the contact rate among onboard crewmembers, latency period, and recovery time – do not change over the course of simulation.

### Sensitivity analysis for population-specific epidemiological parameters

To assess the applicability of these findings to outbreaks with a higher basic reproductive number (R_0_) or to outbreaks with a larger number of initial exposures (E_0_), we varied these parameters accordingly. Both R_0_ and E_0_ are population-specific parameters and are expected to vary by scenario.

## Results

### An SEIR model adequately describes early outbreak dynamics on the Roosevelt

The SEIR model fit to the 14 days of the Roosevelt outbreak data was consistent with a constant contact rate, with strong parameter identifiability for the fit population-specific parameters: the contact period and the initial exposed population (Figure 1B). The fit parameter values and their standard errors (SEs) are summarized in Table 1. The fit contact period is consistent with an R_0_ value of 4.5, which is much lower than the estimate of the R_0_ on the Diamond Princess, which was approximately 15 (Gilday, 2020).

**Table 1:** Model parameters for SEIR model fit to the Roosevelt early disease spread dynamics

| Parameter | Symbol | Value (+/- SE) | Source |
| --- | --- | --- | --- |
| Latency period | A | 3 days | (Lauer et al., 2020) |
| Contact period | B | 2.21 +/- 0.12 days | Model fit |
| Recovery period | C | 10 days | (Rocklöv et al., 2020) |
| Initial exposed population | E<sub>0</sub> | 24.1 +/- 0.80 sailors | Model fit |

### Effectiveness of Roosevelt outbreak control by removal of sailors appears minimally impacted by testing-based enrichment

When we predicted the impact of removal of sailors at random at the reported times, we found that the actual outbreak caseload data falls only slightly below the 95% confidence interval for variation introduced by random evacuation of sailors in a crew of 4,865 (Figure 1C). This suggests that throughout the outbreak interval, the evacuated population was minimally enriched for infected and exposed sailors relative to the population on board the ship – that is, the evacuees were effectively selected at random from the sailors on the ship. This result is surprising given that testing on board the Roosevelt increased from approximately 100 tests per day in March to 500 tests per day in April (Peniston, 2020). The apparent failure of the additional tests to result in more efficient removal of actively infected sailors may be explained by long delays in turnaround of test results (Dyer, 2020), as well as the difficulty in identifying SARS-CoV-2 infectious individuals due to delayed symptom onset and asymptomatic infections (He et al., 2020; Johnson et al., 2020). Prior analyses have demonstrated the importance of rapid test results to decrease time to isolation (Johnson et al., 2020).

### In a successful evacuation, most evacuees avoid infection while those remaining on board fall ill

In Figure 2, we assess the impact of a mass evacuation-based outbreak mitigation strategy, in which testing is absent and sailor removals are exposure-agnostic. Our analysis suggests mass evacuation can be an effective strategy by reducing the size of the population exposed to the pathogen. A successful evacuation preserves the health of the vast majority of sailors removed from the ship, but under the simulated outbreak conditions, nearly all sailors remaining on board become infected. In this vein, the simulation predicts that 4,582 of the Roosevelt sailors (97.8%) would have eventually become infected without the Navy’s interventions (Figure 2B). For a successful evacuation plan, the simulations indicate that above a certain threshold for the rate of evacuation, the outbreak size is only dependent on the final number of sailors remaining on the ship. For the outbreak conditions simulated in our model, this threshold falls around a 6% daily rate of evacuation. This result indicates that mass evacuation-based strategies can be successful with an achievable rate of deboarding.

**Fig 2.**
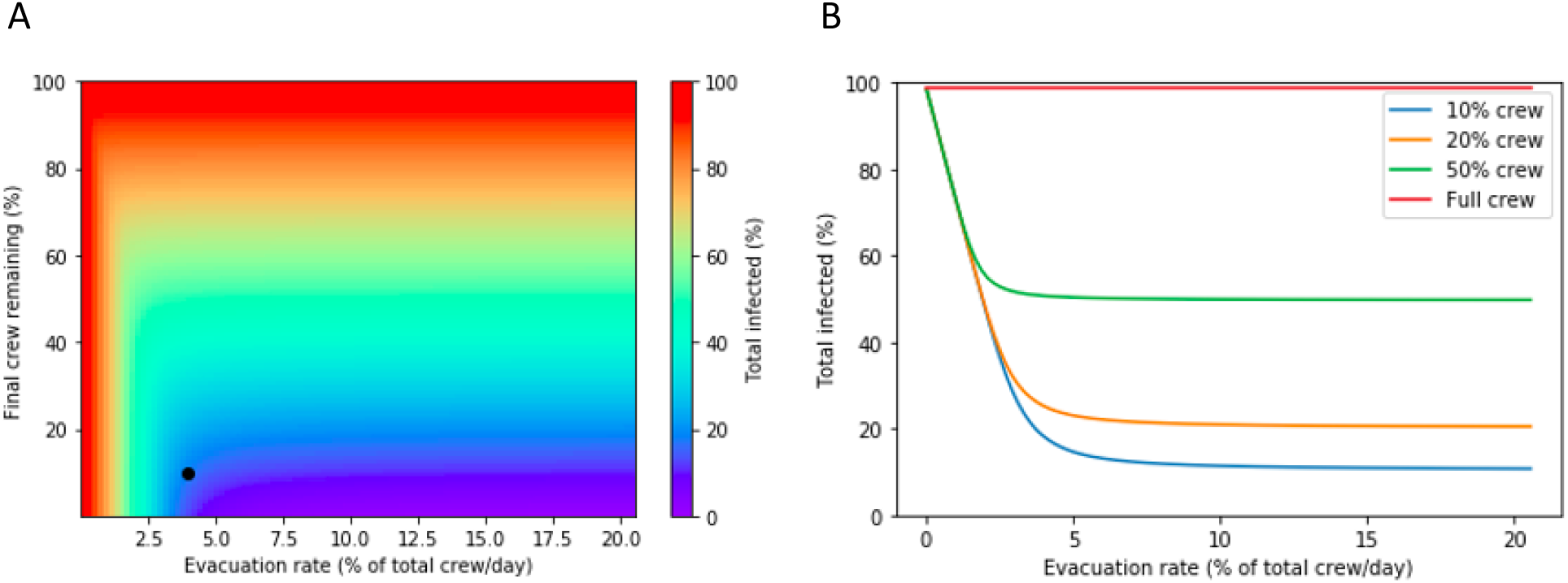
Impact of evacuation strategies on total percentage of initial crew infected. A) Model-predicted extent of outbreak spread under evacuation-based strategies with variable rates and extents of evacuation. Epidemiological parameters are assumed constant throughout the simulation. Black dot represents Capt. Crozier’s proposed strategy, which involved evacuating 4% of the crew daily until 10% remained. B) Percent of crew infected as a function of the evacuation rate for a final crew of 10%, 20%, 50%, or 100% of the initial crew.

In many at-sea scenarios, immediate mass evacuation is not possible until the ship has sailed to port. In these cases, there will be a delay in evacuation after detection of the first case (Figure 3). A delay in implementation reduces the effectiveness of evacuation-based strategies. However, if the delay is not too large, it may be overcome by increasing the rate of evacuation. For example, a one-week delay may be overcome by increasing the evacuation rate from 6% to 10% of the crew daily (Figure 3A). After three weeks, however, the best-case outcomes are no longer accessible, even with a 20% daily evacuation rate (Figure 3B). In agreement with this result, Figure 3C demonstrates that delayed implementation time erodes the effectiveness of even aggressive evacuation strategies.

**Fig 3.**
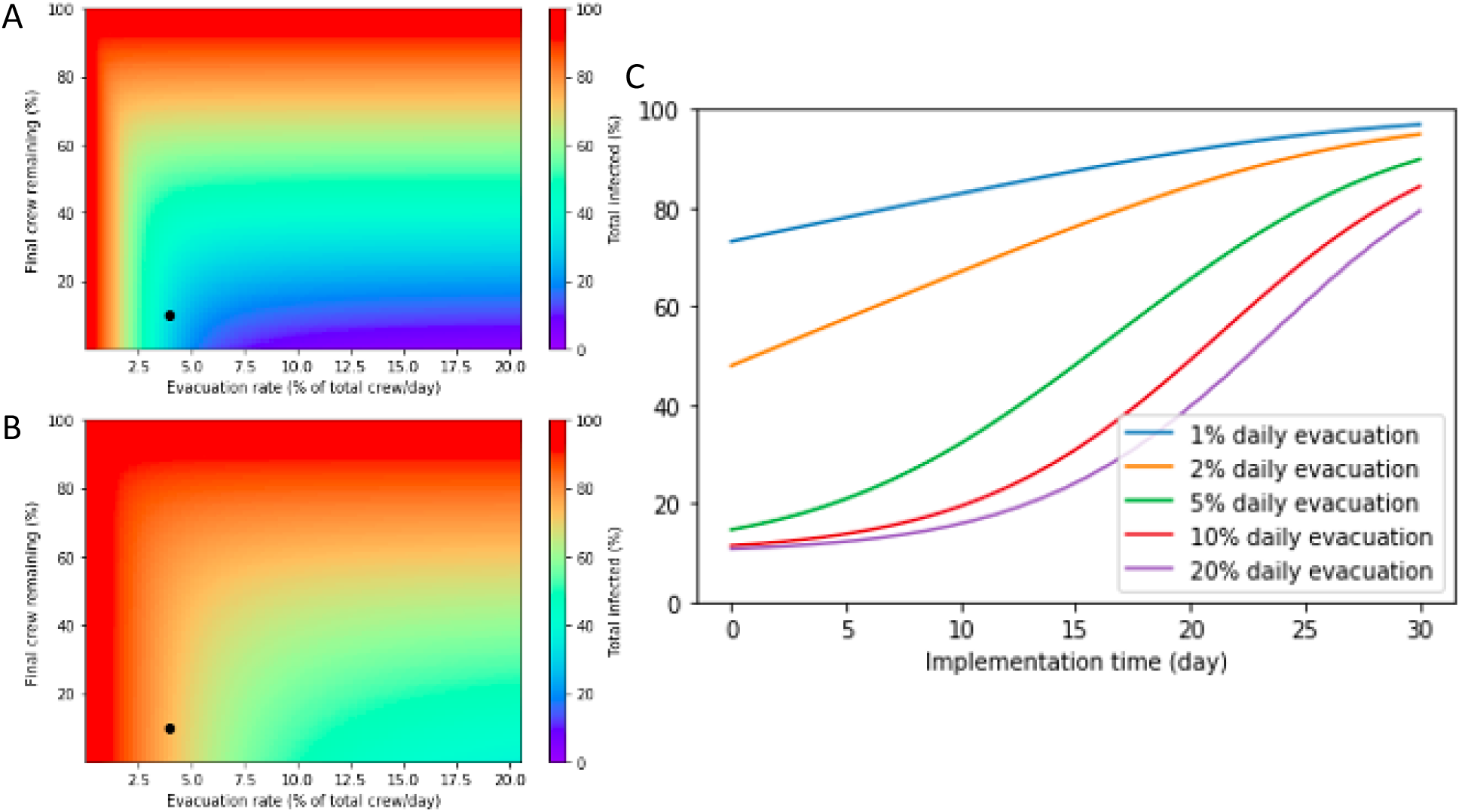
Effectiveness of evacuation depends on speed of implementation. Evacuation strategy implementation on A) day 7 or B) day 21 after detection of the first case. Black dots represent Capt. Crozier’s proposed strategy, which involved evacuating 4% of the crew daily until 10% remained. C) Dependence on time of implementation for retention of a 10% skeleton crew at a 1%, 2%, 5%, and 10% daily evacuation rate.

Capt. Crozier recommended rapid evacuation at a rate of 200 sailors per day (approximately 4% of the initial crew) until 10% of the crew remained to maintain essential functions aboard the Roosevelt. If this strategy had been implemented immediately at detection of the first case, the SEIR model predicts an outbreak size of 733 infected sailors (15%) by May 5 in this scenario, a significant improvement on the actual case count of 1,156 by May 5 (Moriarty, 2020). The downside of this strategy is that the sailors remaining on the ship are intended to perform important and physically demanding tasks, but nearly all of them will become infected, likely impeding their work. Given that data collection stops on May 5 for the Roosevelt, it is unclear what happened to the sailors remaining on board after evacuation.

### Testing allows larger crews to remain safely aboard

Guaranteeing a healthy crew on board a disease-burdened ship is impossible in an evacuation-only response model (Figure 2). Implementing an effective testing-based isolation strategy significantly improves outcomes for crewmembers remaining on board the ship (Figure 4). As shown in Figure 4A, Capt. Crozier’s target crew of 10% of the initial population could have been safely maintained aboard if a testing rate of 5% daily relative to the total crew size (50% of the final crew size) and an evacuation rate of 5% daily had been implemented immediately after detection of the first case. The simulation assumes testing readouts and removal of positive cases are instantaneous and that testing accuracy is perfect. Abbott’s BinaxNOW COVID-19 rapid tests, which recently received emergency authorization from the FDA, may meet these criteria (Koval, 2020). Similar to the evacuation-only case, the rate of evacuation does not significantly impact outbreak outcomes above a certain threshold. In this case, an evacuation rate of approximately 5% daily is required for optimal results. For evacuation rates greater than this threshold, the impact of testing is nearly switch-like. If testing is implemented at a higher than 4% daily rate, the outbreak size is very small (<5%). For lower rates of testing, the outbreak size is a much larger fraction of the total crew.

**Fig 4.**
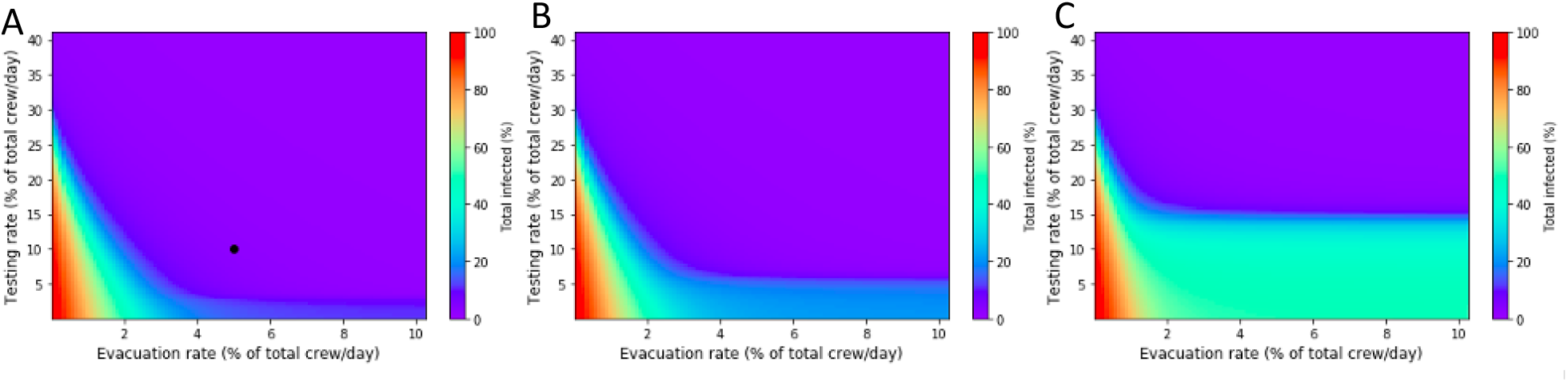
More tests are needed to safely retain a larger crew. Impact of testing and evacuation rates on outbreak size when A) 10%, B) 20%, or C) 50% of the initial crew remains after evacuation. Black point on A) represents the Navy’s intended strategy for 500 tests per day combined with Capt. Crozier’s recommendation for 5% daily evacuation.

The model also demonstrates that retaining a larger crew is feasible. A higher testing rate is required to achieve outbreak containment with a larger crew remaining on board (Figure 4B, C). While a less than 5% rate of daily testing relative to the original crew size can contain disease when 10% of the initial crew remains on board, a daily testing rate of at least 25% is required for containment when 50% of the crew remains on board. As expected, the epidemiological cost of insufficient testing is larger if more crewmembers remain on board. Figure 5 further demonstrates the feasibility of retaining crews of varying sizes if extensive testing is implemented. The steep relationship between testing rate and disease control is readily observed in this figure; a two-fold reduction in testing frequency results in near-complete abolition of testing-driven improvements in outbreak outcome. The full-crew simulations are particularly important to guide outbreak response in scenarios where onboard removal by isolation is possible but mass evacuation to land is not.

**Fig 5:**
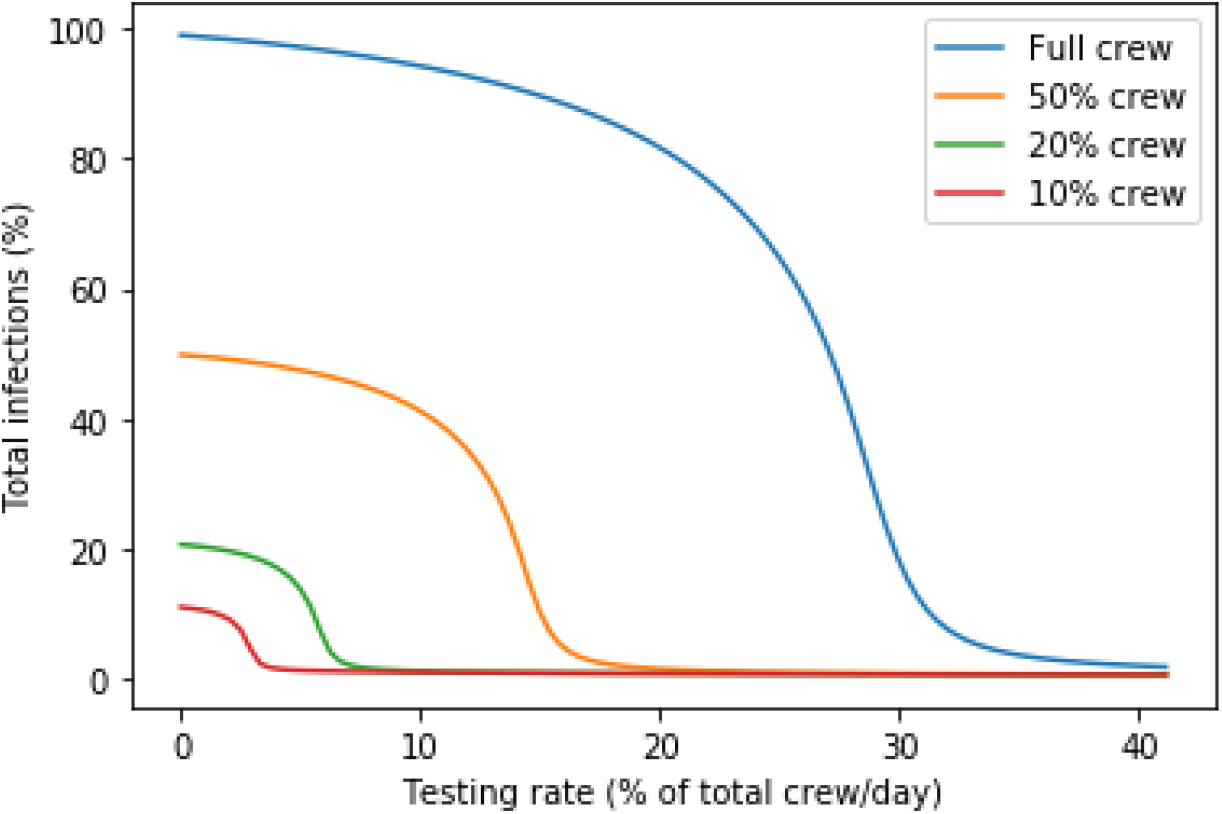
High-frequency testing and targeted isolation results in outbreak containment for various crew sizes. Crewmember evacuation occurs at a rate of 10% daily until the target crew size is reached. Test randomly administered to the crew with isolation of detected cases. Testing rate is relative to the size of the initial crew.

An important caveat to testing-based strategies is that their success relies on the immediate isolation or removal of positive cases. This means that the functional size of the crew may be smaller than the target size due to removal of infected individuals upon detection (Figure S1). The simulation assumes that all detected positives can be removed. If the testing strategy is highly effective, the fraction of the crew removed due to infection is small (<5%). If testing is ineffective, more than 50% of the target crew may require removal upon testing positive. If the testing regime is highly ineffective, the intended number of crew members may remain on duty due to extensive undetected cases.

### Optimal evacuation and testing rates are generally applicable over a range of R_**0**_ **and E**_**0**_

Although recovery time and latency period are disease-specific parameters expected to hold constant across demographically comparable populations, R_0_ and E_0_ are population and outbreak-specific. To assess the applicability of our findings to other at-sea scenarios, we performed a sensitivity analysis to gauge the impact of changes in R_0_ and E_0_ on the effectiveness of a successful outbreak containment strategy. The selected strategy involves evacuation of 5% of crew members daily until 10% of the initial crew remains; testing at a rate of 10% of the total crew daily; and immediate implementation of both measures at the time of detection of the first case (Figure 6). This strategy corresponds to Capt. Crozier’s suggested evacuation plan and the Navy’s intended testing capacity (Gilday, 2020). Our analysis suggests that this strategy is robust to significant increases in R_0_, representing scenarios with higher rates of disease-spreading contact among crewmembers, as well as significant increases in E_0_, representing late detection of the outbreak or a super-spreader disease introduction event.

**Fig 6.**
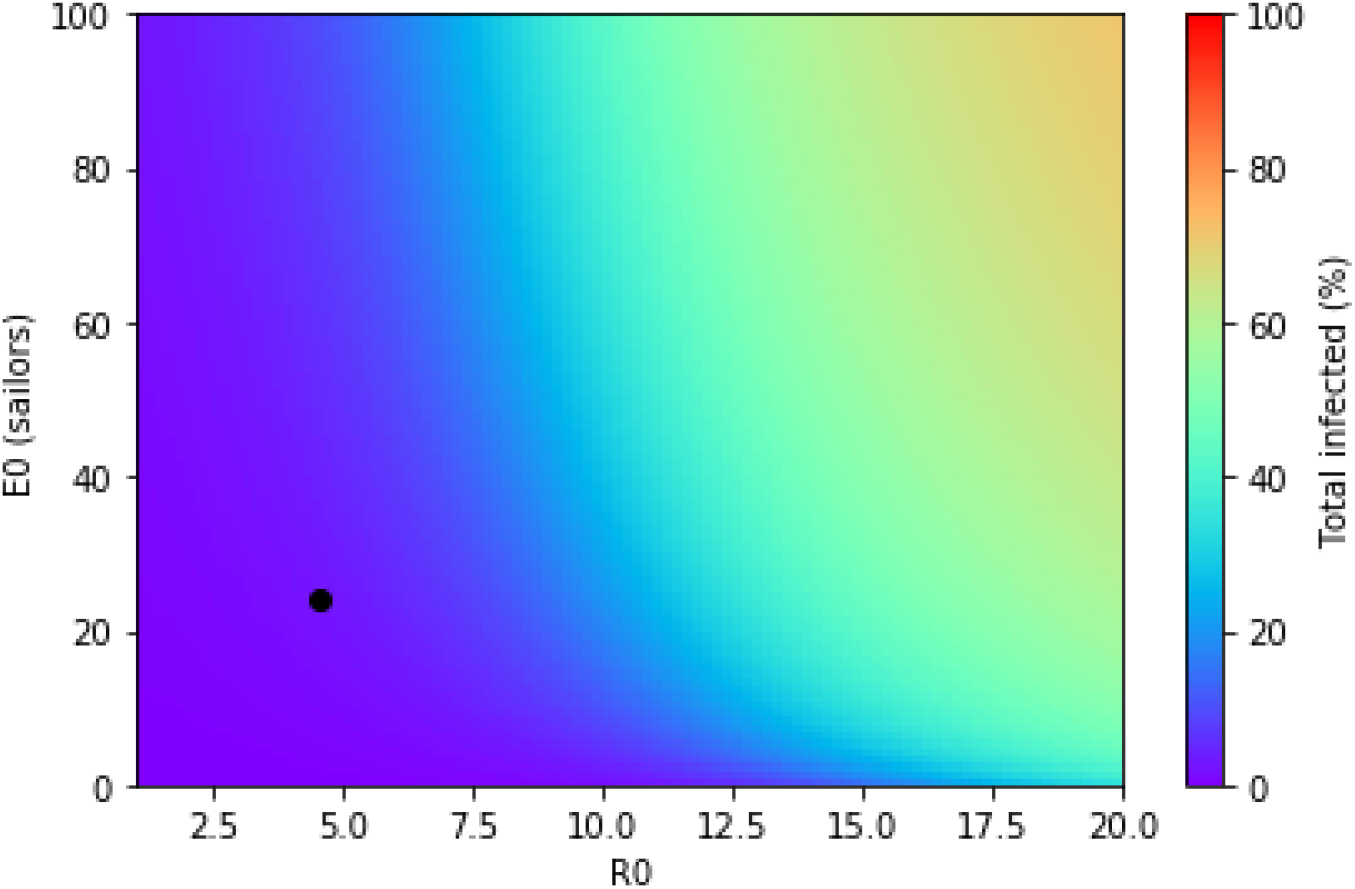
Sensitivity analysis: the Navy’s strategy is successful under a wide range of possible outbreak parameters. In this heatmap, the following strategy is simulated under a variety of disease spread scenarios: after detection of the first case, evacuation of 5% of sailors per day until a crew of 10% remains; testing is carried out at a rate of 10% of the total crew per day. Black point represents fit parameters for the Roosevelt dataset. The purple region represents combinations of R_0_ and E_0_ for which containment is successful.

## Discussion

Disease control in the early stages of a pandemic depends significantly on the ability to contain shipborne outbreaks, as these provide significant opportunities for pathogens to spread in an uncontrolled manner and propagate internationally when passengers are released before the outbreak has run its course. The challenge of managing shipborne disease outbreaks has been evident throughout history: the concept of quarantine itself dates back to restrictions placed on sailors in the era of the Black Death; in 1918, troopships were instrumental in spreading the influenza pandemic (Baraniuk, 2020); and in recent years, the cruise ship industry (“COVID-19 pandemic on cruise ships,” 2020; El Damanhoury and Cullinane, 2019; Hines, 2020) and the Navy (Aquino and Brice, 2014; Khaokham et al., 2013) have both struggled with a variety of infectious disease outbreaks on ships.

A generalized quantitative examination of disease control strategies can thus provide valuable inputs into public health strategy. Our work uses a model-based approach, rooted in the characteristics of the Roosevelt outbreak and based on a counterfactual analysis, to explore the effectiveness of different strategies in shipborne disease control.

Shipborne outbreaks are particularly challenging to control because of the high rate of spread and the ineffectiveness of social distancing and contact tracing in the closed confines of an onboard setting. Reports estimated the R_0_ on the Diamond Princess to be as high as 15 (Rocklöv et al., 2020), with the virus spreading evenly throughout all of the ship’s 18 decks primarily as a result of close-range aerosol transmission (Grasso Macola, 2020). According to another study, the use of social distancing and mask wearing on the Roosevelt had a minimal impact on likelihood of infection (For example, the percentage of sailors infected was 55% vs 80% for mask wearing or not, and 55% vs 70% for social distancing or not (Payne, 2020)).

Faced with these constraints, passenger ships can quickly become large floating reservoirs of disease. This work outlines several practical strategies to avoid this outcome in different scenarios (Table 2). First, in the case of a novel pathogen (at the start of the next pandemic), the best strategy for shipborne outbreaks is likely to be mass evacuation. We note that in the case of the Roosevelt, this was likely made impossible by political constraints. There was a nine-day time lag between the detection of the first case, while the Roosevelt was underway in the Phillipine Sea, and the start of mass evacuation in Guam, which was at most two days’ sail away (Peniston, 2020). Similarly, at the outset of the Diamond Princess outbreak, the ship was quarantined for nearly a month in Yokohama harbor, with her passengers onboard (Grasso Macola, 2020). Delays in mass evacuation at the beginning of a shipborne outbreak can create a much larger problem for local authorities, as was observed in the Diamond Princess outbreak, where passengers released from the ship went on to spread the disease locally (Endo, 2020; Rich and Yamamitsu, 2020). In this context, it is encouraging that a number of cruise lines have proposed agreements with local authorities that expedite rapid evacuation to shore at the start of an outbreak (Sebastian, 2020).

**Table 2:** Recommendations for at-sea epidemic management under different constraints.

| **Epidemic scenario** | **Availability of testing** | **Isolation possible?** | **Mass evacuation possible?** | **Recommendation** |
| --- | --- | --- | --- | --- |
| Novel pathogen | No | Unknown | Yes | Mass evacuation |
| Diamond Princess outbreak (cruise ship) | Yes (limited) | No | Yes | Mass evacuation |
| Roosevelt outbreak (carrier during peacetime) | Yes | Yes (limited) | Yes | Widespread surveillance testing; evacuation of a fraction of crew upon testing shortages or outbreak growth |
| Carrier strike group at war | Yes | Yes (limited) | No | Widespread surveillance testing, isolation of positive cases |
| Submarine | Yes | No | No | Quarantine and testing of full crew before entry |

Recently, the CDC published new guidance to facilitate a return to cruising, stipulating antigen testing of all passengers and social distancing and symptom-based screening of passengers while underway. However, a recent outbreak at the Rose Garden of the White House demonstrated the limitations of antigen tests (Facher, 2020), for which the CDC itself regards negative results as “presumptive” (CDC, 2020). Previous studies on the Roosevelt demonstrated the limitations of social distancing in an onboard setting (Payne, 2020), and our prior work has demonstrated the limited utility of symptom-based testing for the containment of SARS-CoV-2 outbreaks (Johnson et al., 2020). As an alternative, we suggest requiring negative PCR tests for all passengers after a 14-day quarantine, as the CDC requires for crewmembers (HHS, 2020).

Arguably, if testing on the Roosevelt had been operating at full capacity, isolating the infected sailors by airlifting them to land using the organically embedded air wing of the Roosevelt (two C-2 Greyhounds and six MH-64 Sea King helicopters) could have had a significant impact on the growth and spread of the outbreak (“USS *Theodore Roosevelt* (CVN-71),” 2020). Although testing was slow to ramp up in the specific case of the Roosevelt, mass testing and targeted removal can be decisive in enabling continued safe operations. The relationship between testing frequency and strategy effectiveness is steep: low levels of testing are ineffectual, while high levels of testing can be highly effective. For a ship with similar disease spread dynamics as the Roosevelt, a crew of any size can be maintained aboard while containing disease if testing is sufficient. Thus, the availability of widespread and rapid testing is a crucial prerequisite for naval readiness in pandemic settings. A scenario-specific and model-based assessment of the required testing frequency is desirable, as small changes in the effective rate of testing (by reduced test sensitivity, slower turnaround times) may render testing ineffective. It is encouraging that the Navy quickly learned from the Roosevelt outbreak to implement rapid control of a second outbreak on the destroyer USS Kidd using proactive testing-and-isolation strategies, even as the Roosevelt outbreak was still being managed (Olson, 2020).

Finally, when isolation and evacuation are not possible, such as in a submarine, quarantine and testing of the full crew before entry are the only feasible strategies for disease control.

We note that lowering the R0 will also facilitate disease control on ships. A number of authors have pointed out the risk posed by unfiltered forced-air ventilation systems (Baraniuk, 2020; Malone, 2020), and installing higher-quality filters on heating, ventilation and air-conditioning systems is an easy step to take in reducing the rate of transmission of an airborne pathogen. Our work contains a number of simplifying assumptions that are worth exploring in further work. We assumed that testing was perfectly accurate and instantaneous, resulting in an optimistic assessment of the efficacy of testing-based strategies. Further study is required to assess the impact of testing limitations on these strategies and to identify the time to outbreak detection for a given testing frequency. Additionally, our results are dependent on the transmissibility of disease; these parameters will vary in other settings, so the specific recommendations of this work must be updated to apply to other pathogens, although the model-based approach is broadly applicable.

When the next pandemic starts, the ships will still be at sea. It is safe to say that many of the issues experienced at the onset of the COVID-19 pandemic will repeat themselves again in the not-too-distant future. It is our hope that model-based approaches can be used to develop generalizable strategies for disease control, such as those presented here.

## Data Availability

All data used for modeling is derived from publicly available sources.

