## Supplemental Figure for "COVID-19 isolation and containment strategies for ships: Lessons from the USS Theodore Roosevelt outbreak"

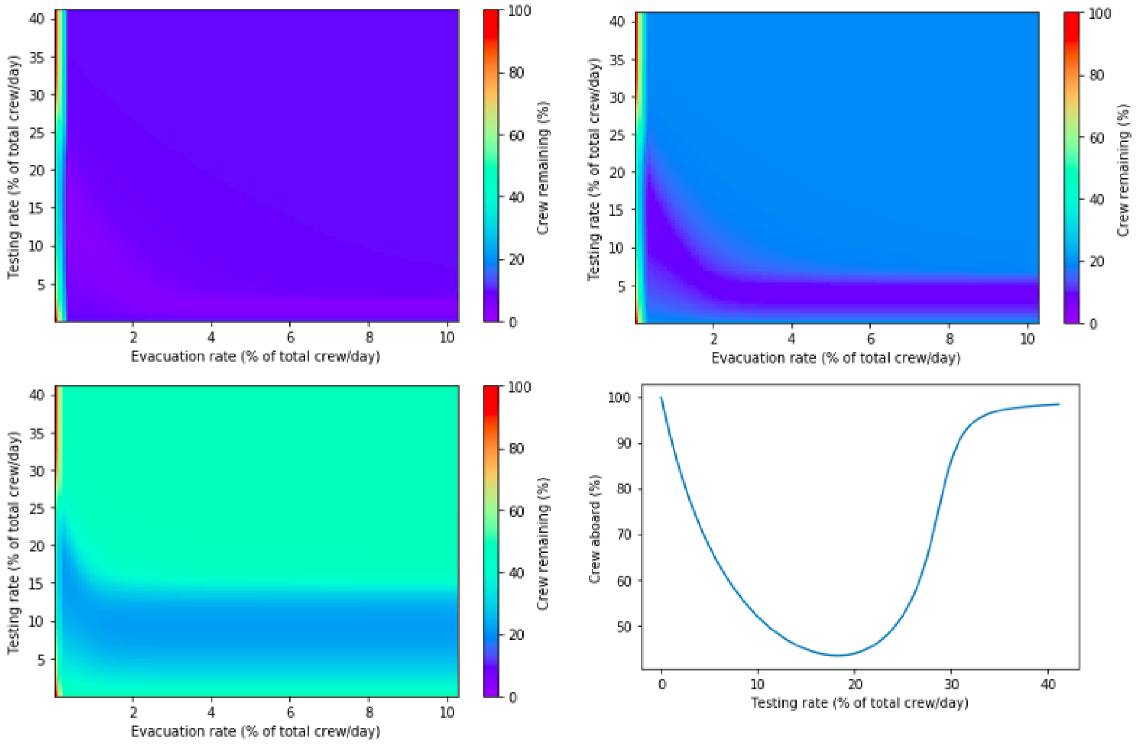


**Fig S1.** Target skeleton crew is unattainable when testing is insufficient or evacuation is too slow. Final crew remaining on board and out of isolation for a target crew of A) 10%, B) 20%, C) 50%, and D) 100% of the initial population.
