## Supplemental Methods for "COVID-19 isolation and containment strategies for ships: Lessons from the USS Theodore Roosevelt outbreak"

### Stochastic adaptation of the SEIR model to predict impact of mass evacuation

We adapted the SEIR model to evaluate the Navy's mass evacuation strategy in the Roosevelt outbreak. To predict the impact of random mass evacuation and compare it to real outcomes, we simulated stochastic removal in which sailors are selected for evacuation at random according to the reported timing and number of sailor evacuations. We simulated stochastic evacuations by implementing a gating criterion for randomly generated decimal values. The gating ensures that the probability of removal from each of the on-board SEIR compartments is proportional to the number of sailors in the compartment at that time. This means that in the simulation, sailors selected for removal were not enriched from infected or exposed categories, as might have been intended. We assume that all evacuations occur at the beginning of the corresponding simulation day. After the evacuated sailors were removed from their respective compartments, we ran the deterministic SEIR model forward until the next set of removals or the end of the simulation. The resulting SEIR dynamics from 10,000 simulations of random removals were used to build 95% confidence intervals for the expected infections over time. Based on these confidence intervals, we compare the observed infections to the expected variability introduced by random evacuations to determine whether actual outcomes were superior to fully random evacuation.

### Deterministic adaptation of the SEIR model to predict impact of mass evacuation and testing-targeted isolation

To assess efficacy of mass evacuation and testing-targeted isolation strategies, we implemented a deterministic ODE-based model accounting for random mass evacuation and testing-based isolation of sailors from the SEIR compartments aboard the ship. The model predicts the total number of infections over the course of the outbreak and the number of healthy sailors aboard the ship over time. An advantage of this deterministic model is its independence of population size, allowing its predictions to be extended to military and passenger ships of all sizes. For the mass evacuation case, the rate of evacuation from each compartment is directly proportional to the compartment's percentage of the on-board population and the ship-wide evacuation rate. This reflects random mass evacuation. We further extended the SEIR model to account for a testing-based rate of removal of the infectious population dependent on the frequency of testing in the population aboard the ship. We assumed that selection for testing among the crew is random and that test results are received instantaneously. To keep the analysis streamlined, we also assume that the test is perfect: all infected crew test positive upon receipt of a test, while susceptible, exposed, or recovered crewmembers test negative. We also assume that all sailors testing positive are immediately removed from the ship, that compliance is perfect, and that individuals removed do not return to work during the simulation interval. Thus, a positive test results in removal from the on-board infectious compartment and a corresponding change in the epidemiologically relevant population on the ship. Tests are administered at the beginning of each simulation day.

In all model implementations, the total number of cases is tracked, including all sailors who become infected while on board the ship and sailors who are exposed on board but do not become infectious until after their evacuation. From a modeling perspective, this means that all compartments are tracked aboard the ship, while only pre-infectious exposed sailors are tracked in an on-shore compartment. We assumed no additional transmission occurred in on-shore isolation facilities. The total number of cases is stored in a separate variable accounting for exposed individuals that develop infections after evacuation.

Equations 6-11: ODEs for SEIR model with testing-based and random removals

$$\frac{dS}{dt} = -\frac{1}{B}SI - \frac{r_{evac}S}{N}$$

$$\frac{dE}{dt} = -\frac{1}{A}E + \frac{1}{B}SI - \frac{r_{evac}E}{N}$$

$$\frac{dI}{dt} = -\frac{1}{C}I + \frac{1}{A}E - \frac{r_{evac}I}{N} - \frac{r_{test}I}{N}$$

$$\frac{dR}{dt} = \frac{1}{C}I - \frac{r_{evac}R}{N}$$

$$\frac{dE_{evac}}{dt} = -\frac{1}{A}E_{evac} + \frac{r_{evac}E}{N}$$

$$\frac{dCases}{dt} = \frac{1}{A}E + \frac{1}{A}E_{evac}$$

Initial conditions for the SEIR model:

$$I(0) = 1/N, E(0) = E_0/N, S(0) = R(0) = 0$$

where $r_{evac}$ is the fraction of the initial crew removed daily, $r_{test}$ is the rate of testing, and N is the total crew on board (equal to the sum of S, E, I, and R, which declines as sailors are removed or evacuated).
